# Disentangling Environmental and Genetic Influences on Associations Between Childhood Bullying Victimization and Psychotic-Like Experiences

**DOI:** 10.64898/2026.02.04.26345591

**Authors:** Nicole R. Karcher, Deanna M. Barch, Hans Oh, Sarah E. Paul, K. Juston Osborne, David AA Baranger, Ryan Bogdan, Arpana Agrawal, Emma C. Johnson

## Abstract

Psychotic-like experiences (PLEs) are common in youth and predict later mental health problems. Bullying victimization is a robust environmental risk factor for psychopathology including PLEs, but whether its association with PLEs reflects shared genetic liability, individual-specific putatively causal effects, or reciprocal processes is unclear. We analyzed seven waves of longitudinal data from the Adolescent Brain Cognitive Development (ABCD) Study, examining associations across the sample in addition to leveraging within-family comparisons among twin and sibling pairs who were concordant or discordant for exposure to bullying victimization. Using linear mixed-effects and cross-lagged models, we found that youth reporting bullying victimization were more likely to endorse significantly distressing PLEs than non-victimized youth (caregiver-reported odds ratio=2.35; youth-reported odds ratio=4.10). Longitudinal analyses revealed bidirectional associations: prior bullying predicted subsequent increases in distressing PLEs, and prior PLEs predicted elevated risk of later bullying victimization. Genetically-informed within-family analyses indicated that both shared genetic influences and individual-specific factors contributed to these associations; critically, bullied youth exhibited higher odds of distressing PLEs than their non-exposed siblings (youth-reported odds ratio=6.67; 95%CI:4.96-8.96), consistent with an individual-specific effect of victimization. Together, these findings suggest that bullying and PLEs are linked through reciprocal developmental processes that are not fully explained by familial confounding. More broadly, our results highlight bullying prevention as a plausible leverage point for reducing early psychosis-spectrum risk and illustrate the value of integrating within-family designs to help disentangle genetic and environmental contributions to mental health outcomes in adolescence.

**Significance Statement:** Understanding how early adversity shapes mental health trajectories is important for science and public policy. Using nationally representative, longitudinal twin and sibling data, analyses show that bullying victimization and psychotic-like experiences in youth are linked through reciprocal processes that cannot be fully explained by shared genetics or family background. Bullied youth were more likely to endorse distressing psychotic-like experiences than their own non-bullied siblings, providing rare evidence for individual-specific effects of bullying victimization. Early psychotic-like experiences also increased subsequent risk of being bullied, suggesting a potential feedback loop that may compound risk. These findings demonstrate how social environments and mental health dynamically interact and point to bullying prevention as a population-level strategy with potential to reduce early psychopathology risk.

---

Psychotic-like experiences (PLEs) can include perceptual distortions and unusual thought content and are relatively common (e.g., 17% lifetime prevalence in U.S. youth).(1–4) While not all PLEs are clinically relevant, 43-46% of youth PLE with PLE report that the experiences are distressing,(5, 6) and 57-92% of those with significant distressing PLEs develop a diagnosable mental health problem later in adulthood, notably increased risk for psychosis and other serious mental health concerns.(7, 8) Models of worsening PLEs indicate that symptoms may increase due to factors including accumulating alteration in pathophysiology, genetic propensity, and due to exposure to environmental insults, including bullying victimization.(9–11)

Childhood and adolescent psychopathology, including PLEs, have been consistently linked to experiences of bullying victimization.(12–18) Longitudinal research indicates that bullying victimization experiences potentially increase the risk for later development of psychotic symptoms and may play a causal role in the emergence of psychosis in vulnerable individuals.(17) Bullying victimization may be specifically linked to PLEs, perhaps even over and above other types of psychopathology. For example, bullying victimization involves being the recipient of intentional harm and often involves social threat, which can contribute to the development of PLEs, including suspiciousness, among those victimized.(19) Experiences of bullying victimization can also promote feelings of social defeat, or feelings of being an outsider, which are additionally implicated in the pathogenesis of psychosis.(20) Conversely, the experience of PLEs may lead to being perceived as odd, which may lead to further bullying victimization.(17, 21) While there is evidence that bullying victimization and PLEs are linked, information on the directionality of associations is lacking and potentially confounded by shared genetic predisposition. However, with the advent of large-scale prospective longitudinal studies such as the Adolescent Brain Cognitive Development (ABCD) Study, we now have a rare opportunity to understand the bidirectional relationships between bullying victimization and PLEs. Further, the availability of genetically related individuals, including twins and non-twin siblings, provides opportunities to parse the role of shared genetic predisposition that might influence PLEs and elevate likelihood of experiencing bullying victimization from individual-specific and potentially causal effects of bullying victimization on PLEs (or vice versa).

The present manuscript leveraged the ABCD Study to test contrasting hypotheses about the link between bullying victimization and PLEs. Specifically, we examined whether experiencing bullying victimization contributes to increased PLEs, consistent with an individual-specific or potentially causal environmental effect, versus whether both bullying victimization and PLEs arise from shared underlying vulnerabilities, such as genetic predisposition. First, we tested in the overall sample whether ever experiencing bullying victimization was associated with ever experiencing significantly distressing PLEs across seven waves of ABCD data. Second, we examined leading and lagging associations between bullying victimization and PLEs to examine whether significantly distressing PLEs increased following the experience of bullying victimization or vice versa. Third, we conducted within-family comparisons using twin and sibling pairs, including those discordant for bullying victimization exposure (and separate analyses examining youth discordant for PLEs), to further disentangle individual-specific (potentially causal) effects from shared familial influences. As twin and sibling pairs share at least 50% of genetic material, any excess presence of PLEs in the sibling exposed to bullying relative to their unexposed co-sibling may be viewed as evidence in favor of putatively causal individual-specific influences. These analyses provide a critical test of whether the observed association reflects environmentally driven risk, shared heritable liability, or both, clarifying the role of the environment related to and generated by experiencing bullying victimization in worsening youth PLEs.

## METHODS

### Participants

The ABCD Study is a large-scale study of adolescent health and development that recruited and is longitudinally following 9-10-years-olds from 21 research sites (https://abcdstudy.org/study-sites/) across the United States. The ABCD study aimed to recruit a sample reflecting the demographic variation of the U.S. population, recruiting children using probability sampling from both public and private elementary schools. Study-wide exclusionary criteria were as follows: child not fluent in English, MRI contraindication (e.g., irremovable ferromagnetic implants or dental appliances, claustrophobia, pregnant), major neurological disorder, gestational age less than 28 weeks or birthweight less than 1,200 grams, history of traumatic brain injury, or had a current diagnosis of schizophrenia, autism spectrum disorder (moderate, severe), mental retardation/intellectual disability, or alcohol/substance use disorder. (22–24) Parents provided written informed consent and all children provided assent.

Data were drawn from ABCD Data Release 6.0 (https://doi.org/10.82525/jy7n-g441), which includes 7 waves of data: baseline (N=11,868; mean age=9.94(SD=0.63)), 1-year follow-up (N=11,219; mean age=10.96(SD=0.64)), 2-year follow-up (N=10,973; mean age=12.07(SD=0.67)), 3-year follow-up (N=10,450; mean age=12.96(SD=0.65)), 4-year follow-up (N=9,739; mean age=14.17(SD=0.71)), 5-year follow-up (N=8,885; mean age=15.07(SD=0.67)), and a portion of 6-year follow-up (N=5,056; mean age=16.07(SD=0.66)); Table 1 for sample characteristics).

**Table 1.** Whole Sample (n=11,868) Characteristics.

| Variable | Frequency | Mean/Proportion | SE |
| --- | --- | --- | --- |
| Age at baseline (in years) | 11868 | 9.96 | 0.01 |
| Sex (% Female) | 5678 | 47.80% | 0.005 |
| Caregiver-Reported Bullying |  |  |  |
| Baseline | 1814 | 15.30% | 0.003 |
| 1-Year | 1860 | 15.70% | 0.003 |
| 2-Year | 1481 | 12.50% | 0.003 |
| 3-Year | 945 | 8.00% | 0.002 |
| Youth-Reported Bullying |  |  |  |
| Baseline | 2084 | 17.60% | 0.003 |
| 1-Year | 672 | 5.70% | 0.002 |
| 2-Year | 135 | 1.10% | 0.001 |
| Significantly Distressing PLEs |  |  |  |
| Baseline | 3157 | 26.60% | 0.004 |
| 1-Year | 2219 | 18.70% | 0.004 |
| 2-Year | 1732 | 14.60% | 0.003 |
| 3-Year | 1446 | 12.20% | 0.003 |
| 4-Year | 1238 | 10.40% | 0.003 |
| 5-Year | 1050 | 8.80% | 0.003 |
| 6-Year | 471 | 4.00% | 0.002 |

Abbreviations. SE=standard error; PLEs=psychotic-like experiences.

### MeasuresProdromal Questionnaire-Brief Child Version (PQ-BC)

Participants completed the previously validated 21-item Prodromal Questionnaire-Brief Child Version (PQ-BC)(5) which asks participants about PLEs and their associated distress. Consistent with previous research,(5, 25) a continuous distressing PLE score for each timepoint (baseline through 6-year follow-up) was formed by summing distress ratings [i.e., 0=no, 1=yes (but no distress), 2-6=yes (1+score on distress scale); range:0-126] for all 21 items. We also created a significantly distressing PLE dichotomous group that defined individuals who endorsed one or more PLE item as significantly distressing (i.e., rating a PLE ≥3 on a five-point scale of distress, score=1)(26) and those endorsed 0 significantly distressing PLEs (score=0) at each timepoint. Follow-up analyses examined whether associations with PLEs were specific to a PLE symptom type, examining summations of significant distress scores for a) suspiciousness, b) unusual thought content, c) perceptual distortions, and d) disorganization items (see Supplemental Table 1 for a mapping of individual items to domains).(27)

### Experience of Bullying Victimization

Caregivers (ABCD Data Release 6.0 contained all data from baseline [n=11,873], 1-year [n=11,219], 2-year [n=10,924], and 3-year [n=10,263] follow-ups) and youth (ABCD Data Release 6.0 contained a subset of data from baseline [n=11,687], 1-year [n=6,534] and 2-year [n=1,373] follow-ups) responded to a single yes/no question as to whether bullying occurred at school or in the neighborhood within the background section of the Kiddie-Structured Assessment for Affective Disorders and Schizophrenia (KSADS) for DSM-5.(28, 29) Caregiver and child-reported bullying victimization were defined separately as the presence of bullying at any time point (n_caregiver_=3802 [32%]; n_child_=2503 [21%]).

For cross-lagged panel analyses, all available data were used, except for youth-reported bullying victimization, in which models focused on baseline and 1-year follow-up, since there were model convergence issues due to low endorsement of bullying amongst available data at 2-year follow-up (n=135; N=1,373).

Follow-up analyses examined a youth-reported question about whether youth were ever the victim of cyberbullying using one question from the Cyberbullying Scale,(30) administered at 2-year follow-up through 4-year follow-up. Separate analyses also examined whether youth endorsed any of the 9 reputational, relational, or overt peer victimization items on the Peer Experiences Questionnaire,(31) which was administered at 2-year follow-up through 6-year follow-up, with analyses focusing on whether youth reported either 1) ever experienced peer victimization or 2) regularly experience peer victimization (i.e., either about once a week or a few times a week).

### Brief Problem Monitor – Youth Form (BPM-Y)

To examine whether associations were specific to PLEs or generalized to other types of youth-reported psychopathology related to bullying victimization, we also examined the BPM-Y internalizing subscale. This 19-item self-report questionnaire includes an internalizing subscale.(32) Responses are recorded on a 3-point Likert scale (0 = Not True; 1 = Somewhat True; 2 = Very True; 777 = Refuse to answer; 999 = Don’t know) and higher scores indicate higher symptomatology. The raw scaled scores produced for internalizing scores were used in the follow-up analyses. To be consistent with PLE analyses, analyses examined whether youth had ever endorsed at least 1 symptom of internalizing symptoms (i.e., endorsed at least one symptom as 2= very true) from 1-year through 6-year follow-up.

### Statistical Analyses

Unless otherwise specified, analyses used mixed-effect models conducted in R using the lme4 package, nesting family and site. Analyses included age and sex as covariates. First, mixed effect models examined associations between ever experiencing bullying victimization and ever experiencing significantly distressing PLEs. Analyses examined whether individuals endorsed at least 1 significantly distressing PLE at any point from baseline through 6-year follow-up (i.e., ‘Ever Endorsed Significantly Distressing PLEs’; n=5,555, [47%]). Follow-up analyses also examined whether findings remained consistent when: 1) including internalizing symptoms in these models, 2) examining alternative youth-reported bullying victimization definitions (e.g., cyberbullying, or either ever experienced peer victimization or regularly experience peer victimization according to the Peer Experiences Questionnaire), or 3) examining type of PLEs, separately examining significantly distressing PLEs summations for suspiciousness, unusual thought content, perceptual distortions, and disorganization items.

Second, reciprocal within-person associations between bullying victimization and psychotic-like experiences (PLEs) were evaluated using random intercept cross-lagged panel model (CLPM; using the lavaan package in R)(33), using timepoints when sufficient data for both PLEs and bullying victimization were available (caregiver-report: baseline through 3-year follow-up, youth-report: baseline and 1-year follow-up). This analysis examined whether bullying victimization reports at each assessment wave predicted PLEs at the next assessment wave, as well as whether PLEs at each assessment wave predicted bullying victimization reports at the next assessment wave (i.e., testing leading and lagging effects of bullying victimization and PLEs). For youth-reported bullying victimization models, we examined a standard CLPM (i.e., not a random intercept CLPM), due to the fewer waves of youth-reported bullying victimization data.

Third, all possible same-sex twin/sibling pairs were drawn from the data (N pairs based on caregiver data:1624; MZ pairs=419; DZ pairs=641; sibling pairs=564; N pairs based on youth data:1845; MZ pairs=419; DZ pairs=641; sibling pairs=785). Twin/sibling pairs were assigned to 4 groups based on exposure to bullying victimization: concordant unexposed (caregiver n =2335; youth n=2656), concordant exposed (caregiver n =356; youth n=204), exposed individuals from discordant pairs (caregiver n=278; youth n=266), and unexposed individuals from discordant pairs (caregiver n=278; youth n=266).(34, 35)

Next, we used Helmert contrast coding to conduct sibling analyses by bullying victimization exposure, examining the relationship between experiences of bullying victimization (i.e., Ever Experienced Bullying) and PLEs (both Ever Endorsed Significantly Distressing PLEs, capturing lifetime endorsement of significantly distressing PLEs, and Significantly Distressing PLEs slope from baseline through 6-year follow-up, reflecting the change in significantly distressing PLEs over time; see Supplemental Figure 1 for a depiction of the Helmert contrast coding).(34) Separate analyses also examine youth discordant for significantly distressing PLEs. Analyses used mixed-effect models conducted in R using the lme4 package, nesting family, site, and twin pair. Three hypotheses were tested: 1) causal (i.e., bullying victimization and PLEs are associated via person-specific, potentially causal factors), by testing whether bullying victimization exposed twins/siblings from discordant pairs differed in PLE scores from their unexposed co-twin/sibling. 2) predispositional (i.e., due to factors shared by members of twin/sibling pairs, including segregating genetic loci), by testing whether the unexposed pairs differed from both members of discordant pairs and concordantly exposed pairs; and 3) graded liability, a variation of the predispositional model (i.e., exposure does not lead to changes in PLEs within discordant pairs), testing whether there is a gradation of liability for PLEs with concordant exposed pairs showing greater liability than discordant pairs. Importantly, these contrasts allowed for the examination of support for all three hypotheses, as the likelihood of causal and correlated liabilities are not mutually exclusive. All models also examined an interaction between MZ twin status with each of the Helmert contrasts, to determine whether associations generalized to MZ twin pairs. Analyses were false-discovery rate corrected across the 6 tested contrasts for caregiver-reported bullying victimization (i.e., causal, predispositional, and graded with either Ever Endorsed Significantly Distressing PLEs and Significantly Distressing PLEs slope, for 6 total contrasts), and across the 6 tested contrasts for youth-reported bullying victimization. Descriptive analyses examined mean differences in PLE scores amongst discordant pairs and amongst MZ twins to examine whether PLEs were higher in the exposed twin/sibling relative to their genetically related co-twin/sibling.

## RESULTS

### Associations between Ever Experiencing Bullying Victimization and Ever Endorsing Significantly Distressing PLEs

See Table 1 for sample characteristics. Mixed-effects models first examined associations between ever experiencing bullying victimization (based on caregiver or youth report) and ever experiencing a significantly distressing PLE across all assessment timepoints. Caregiver-reported and youth-reported exposure to bullying victimization were associated with increased likelihood of endorsing a significantly distressing PLE, especially for youth-reported bullying (Figure 1; caregiver-reported: odds ratio=2.35 (95%CI=2.15,2.56); youth-reported: odds ratio=4.10 (95%CI=3.68,4.58)).

**Figure 1.**
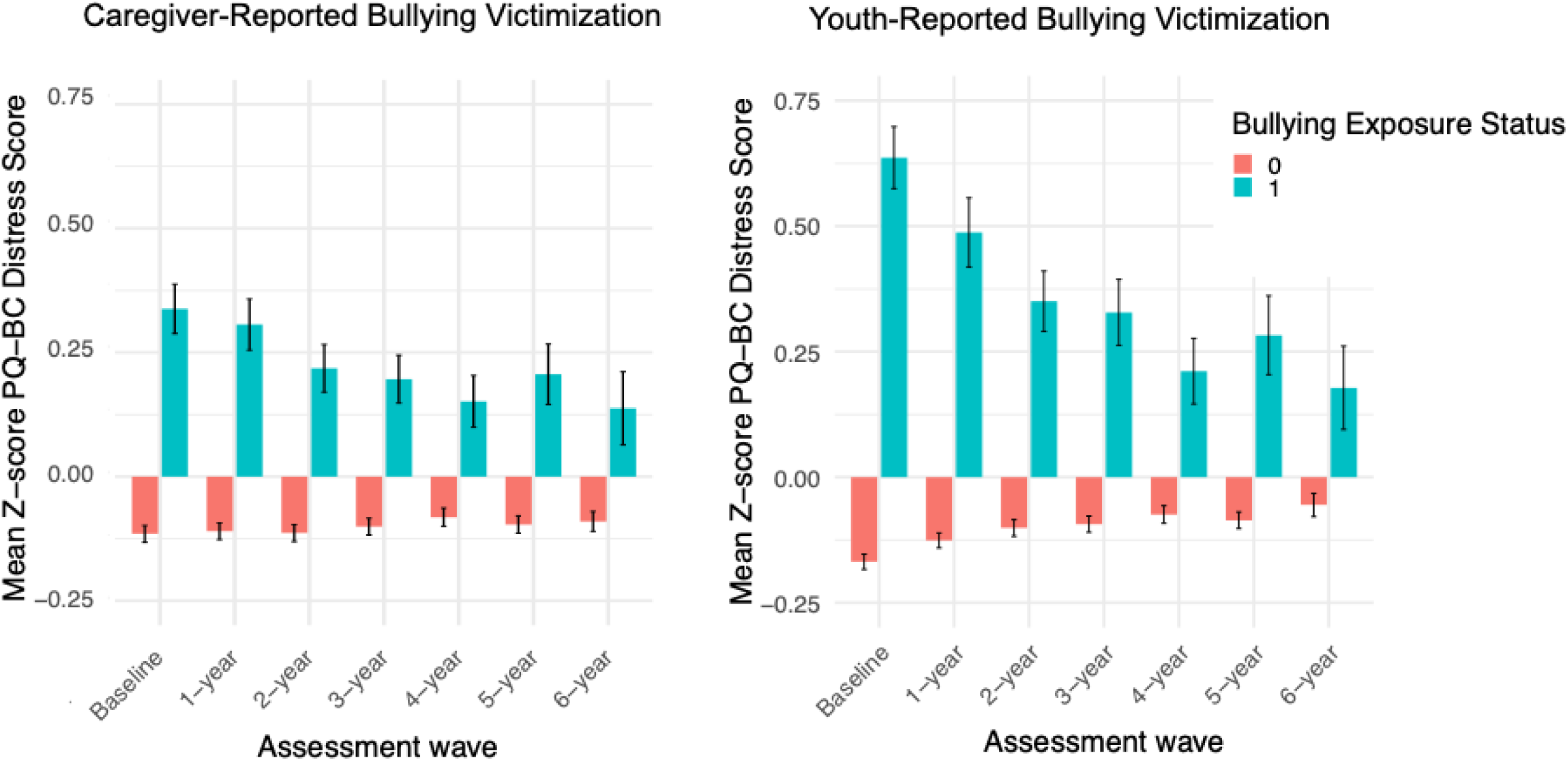
PLE Distress Scores by Caregiver- and Youth-Reported Exposure to Building Victimization. Mean standardized Prodromal Questionnaire-Brief Child version (PQ-BC) distressing psychotic-like experiences (PLEs) score and standard error for each assessment wave by exposure status (0=ever not exposed to bullying victimization, 1=ever exposed to bullying victimization).

Follow-up analyses revealed that associations between PLEs and bullying victimization were consistent across types of bullying victimization (e.g., cyberbullying [odds ratio=3.15, 95%CI=2.85,3.48], or either ever experiencing peer victimization [odds ratio=2.19, 95%CI=1.93,2.48] or regularly experiencing peer victimization [odds ratio=3.34, 95%CI=2.98,3.73]). Associations between bullying victimization with PLEs were also consistent across symptom type, with evidence that bullying victimization was associated with suspiciousness (odds ratios>2.45), unusual thought content (odds ratios>2.08), perceptual distortions (odds ratios>2.23), and disorganization significantly distressing PLEs (odds ratios>2.81).

Lastly, follow-up analyses revealed that the association between bullying victimization and PLEs remained when accounting for internalizing psychopathology. While caregiver-reported internalizing symptoms also predicted bullying victimization (caregiver-reported: odds ratio=1.22 (95%CI=1.05,1.43); youth-reported: odds ratio=1.00 (95%CI=0.90,1.12)), the association between PLEs with bullying victimization was not strongly affected by the inclusion of internalizing symptoms in the model. Odds ratios for the association between bullying victimization with significantly distressing PLEs remained relatively stable after accounting for internalizing symptoms (caregiver-reported: odds ratio=2.34 (95%CI=2.13,2.57); youth-reported: odds ratio=4.22 (95%CI=3.75,4.75)), providing an indication that the association is not driven by comorbid internalizing psychopathology.

### Cross Lagged Associations Between Bullying Victimization and PLEs over Time

Next, random intercept cross-lagged panel models (CLPM) examined leading and lagging effects, asking whether effects were stronger for bullying victimization with later PLEs versus PLEs with later bullying victimization while controlling for concurrent associations. For caregiver-reported bullying victimization, there was strong evidence for bullying victimization relating to later significantly PLEs (from 1-year follow-up through 3-year follow-up: bs>0.043, Zs>4.67, ps<.001), as well as PLEs relating to later bullying victimization (from 1-year follow-up through 3-year follow-up: bs>0.047, Zs>5.84, ps<.001). These same associations were found when examining youth-reported bullying victimization relating to later PLEs at 1-year follow-up, b=0.13, Z=10.81, p<.001, as well as PLEs relating to later youth-reported bullying victimization at 1-year follow-up, b=0.06, Z=7.10, p<.001).

When examining the subset of youth belonging to a twin/sibling pair, for youth-reported bullying victimization, as with caregiver-reported bullying victimization, there was strong evidence for baseline bullying victimization relating to later PLEs at 1-year follow-up (youth-reported: b=0.16, Z=11.62, p<.001; caregiver-reported: b=0.074, Z=3.84, p<.001), however, compared to caregiver-reported bullying victimization there was weaker evidence for significantly distressing PLEs relating to later youth-reported bullying victimization at 1-year follow-up (youth-reported b=0.046, Z=1.37, p=.17; caregiver-reported: b=0.038, Z=2.81, p=.005).

### Twin/Sibling Pair Analyses of the Effect of Ever Experienced Bullying Victimization

There was a robust main effect of bullying victimization exposure, whereby youth exposed to bullying victimization (i.e., exposed members of discordant pairs and members of concordant exposed pairs) were more likely to report having ever experienced a significantly distressing PLE relative to unexposed youth (i.e., members from discordant and concordant unexposed pairs; caregiver-reported: odds ratio=2.06, 95%CI: 1.38,3.07; youth-reported: odds ratio=6.67, 95%CI: 4.96,8.96; Figure 1). Using Helmert contrast coding to test causal, predispositional, and graded liability contrasts (see Supplemental Figure 1 for a graphical depiction), analyses examining sibling pairs organized by caregiver-reported bullying victimization predicting PLEs (i.e., either ever endorsed a significantly distressing PLEs or PLEs slope, see Table 2) found that there was support for the causal (β=0.062, *p*<.001, 95%CI: 0.024,0.099) and graded (β=0.140, *p*<.001, 95%CI: 0.068,0.213) liability contrasts. There was not strong evidence for the predispositional contrast (β=0.004, *p*=.26, 95%CI: -0.024,0.032). There was not strong evidence for interactions with MZ twin status (*Z*s<-0.954, *p*s>.34), indicating that MZ twins showed similar findings to non-MZ twins.

**Table 2.**
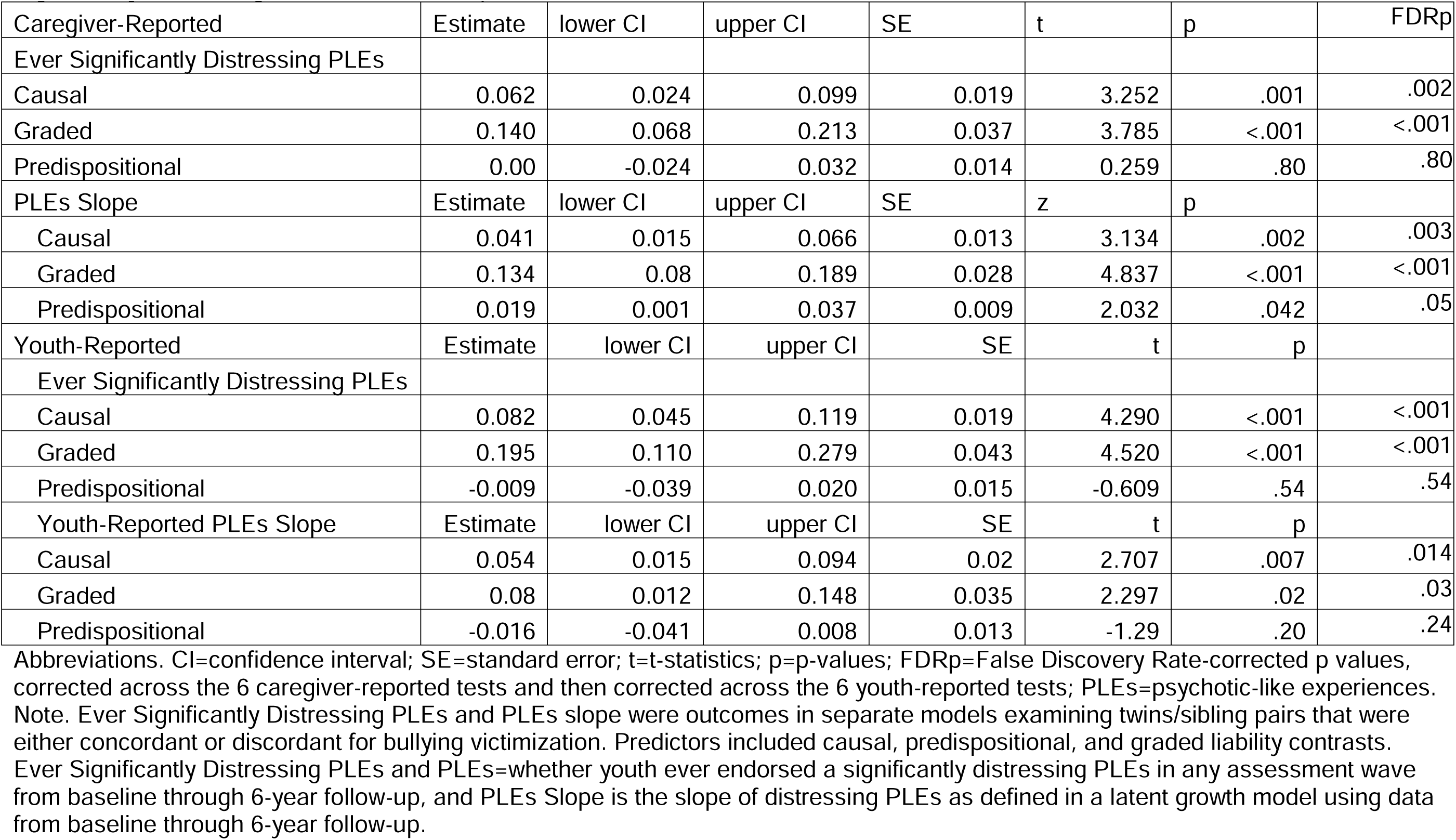
Model Estimates for Causal, Graded, and Predispositional Models Caregiver- and Youth-Reported Ever Experiencing Significantly Distressing PLEs and PLEs Slope.

Findings were consistent when examining youth-reported bullying victimization (see Figure 2 for PLEs by sibling pair type), although when examining youth-reported bullying victimization, there was evidence that support of the causal contrast was attenuated amongst MZ twins, as supported by a MZ x causal interaction, β=-0.049, *p*=.01, 95%CI:- 0.086,-0.012. Regardless, when only examining MZ twins, there was still an exposure effect, whereby MZ twins exposed to bullying victimization showed elevated PLEs compared to non-exposed MZ twins (β=0.282, *p*<.001, 95%CI: 0.203,0.362).

**Figure 2.**
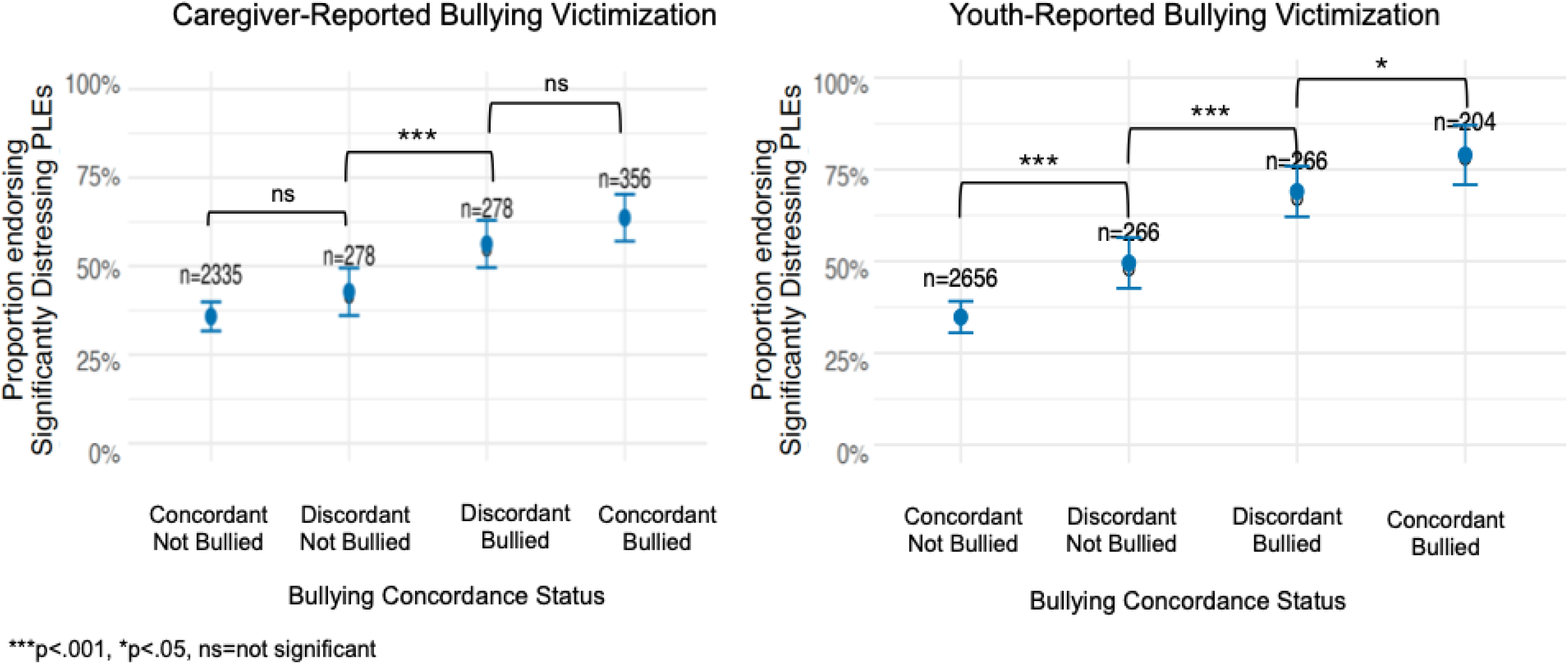
Caregiver- and Youth Reports of Ever Reporting Bullying Victimization by Twin/Sibling Concordance Type. Proportion (and standard error) ever endorsing significantly distressing psychotic-like experiences (PLEs) among each of the bullying victimization concordance types (ie, concordant exposed, discordant exposed, discordant unexposed, concordant unexposed) for twin/sibling pairs. Brackets indicate False Discovery Rate (FDR)-corrected pairwise comparisons shown for selected contrasts to aid visual clarity; all additional pairwise comparisons not depicted were also significant after FDR correction (*p*<.001).”

Analyses examining sibling pairs organized by endorsement of PLEs (i.e., whether youth ever experienced a significantly distress PLEs at any timepoint) predicting caregiver-reported ever experiencing bullying victimization also found support for the causal (β=0.050, *p*<.001, 95%CI: 0.023,0.076) and graded contrasts (β=0.180, *p*<.001, 95%CI: 0.125,0.235; see Supplemental Figure 2), but not strong support for the predispositional contrast (β=0.020, *p*=.07, 95%CI: -0.002,0.043).

## DISCUSSION

The present study leveraged the large-scale, longitudinal ABCD Study to investigate the relationship between bullying victimization and PLEs among youth. Our findings indicate a significant association between bullying victimization and increased PLEs, with robust evidence suggesting that the relationship is influenced by both a potential direct, putatively “environmental” effect of bullying victimization on PLEs as well as graded evidence for the role of shared genetic/familial predisposition of bullying on PLEs (but also of PLEs on bullying). The associations were strongest for youth-reported indices of bullying victimization, and generalized across forms of bullying (e.g., cyber, relational, and reputational peer victimization) and across types of PLEs (i.e., unusual thought content, perceptual abnormalities, suspiciousness, and disorganized speech). There were bidirectional relationships between bullying and PLEs, with evidence for baseline bullying victimization being associated with greater later PLEs but also the reverse. These bidirectional findings underscore the importance of adopting a longitudinal perspective and highlight that bullying victimization might be a potential critical modifiable factor to target in PLE interventions while also encouraging vigilance and support regarding bullying victimization in youth experiencing PLEs.

For youth, reporting bullying victimization was associated with 4.22 times greater odds of endorsing significantly distressing PLEs. This is consistent with and builds upon past research finding a link between bullying victimization and psychosis spectrum symptoms. (12–15, 17, 36, 37) Longitudinal analyses revealed that experiencing bullying victimization was associated with increased PLEs over time and that baseline reports of bullying victimization were significantly associated with later increased distressing PLEs, even after controlling for concurrent associations between bullying and PLEs. These observations align with the social defeat hypothesis,(20) which posits that stressful experiences of exclusion and threat such as those induced by bullying victimization may contribute to dopaminergic alterations and/or other pathophysiological sequelae of acute stress, elevating risk of psychotic symptoms(38–43). However, findings also indicated that those endorsing significantly distressing PLEs are more likely to later endorse bullying victimization. This may be in part because those youth endorsing significantly distressing PLEs are viewed as different or unusual compared to their peers, which results in them preferentially being targeted. Unfortunately, this peer victimization may then later contribute to further worsening PLEs thus perpetuating the cyclical process of stress begetting psychopathology and vice versa. Interestingly, when analyses were isolated to within-families, there was stronger evidence for youth-reported bullying victimization leading to PLEs compared to evidence for PLEs leading to bullying victimization. Further, the association between bullying and PLEs held when incorporating current internalizing symptoms, as well as when examining different types of PLEs (i.e., suspiciousness, unusual thought content, perceptual distortions, or disorganized speech), underscoring that these associations are not solely the result of other types or psychopathology (i.e., internalizing symptoms) or one type of PLEs (i.e., suspiciousness). These analyses extend previous work done in adults and in clinical samples (17) to uncover compelling evidence for bi-directional relationships between bullying and PLEs and adolescence in a non-clinical sample.

Our sibling and twin pair analyses provide nuanced insights into the association between bullying victimization and PLEs. Support for the causal hypothesis was evidenced by higher PLE scores in bullying-exposed members of discordant twin pairs compared to their unexposed co-twin. This suggests that exposure to bullying victimization has person-specific effects, potentially exacerbating PLEs. Even within pairs of individuals that share genetic background and familial environments (i.e., twins and sibling pairs), there was evidence that bullying victimization was linked to greater PLEs. For caregiver-reported bullying victimization, MZ twins showed patterns similar to those observed in non-MZ twin pairs. In contrast, for youth-reported bullying victimization, the causal contrast was attenuated among MZ twins, suggesting weaker evidence for a causal association. Nonetheless, MZ twins who reported being bullied still exhibited higher levels of PLEs than their co-twins who did not report bullying. Together, these findings suggest that bullying victimization may exert individual-specific effects on PLEs beyond shared genetic and familial influences, while also highlighting reporter-specific variability in the strength of individual-specific inference.

Although it is not possible in the current data to rule out all of those influential factors (e.g., the role of stress responsivity), the analyses do add credibility to the hypothesis that differences in significantly distressing PLE endorsement are partly attributable to exposure to bullying victimization. We note that there is evidence for bidirectionality. Potential causal pathways from bullying victimization to PLEs may be related to the effects of increased stress which in turn may lead to greater PLEs.(38) Simultaneously, the predispositional model was also supported, such that even when an individual did not themselves experience bullying victimization, their likelihood of PLEs was heightened if their co-twin/sibling had experienced victimization. While the findings rule out predisposition as the sole driver of the association between bullying victimization and PLEs, it is possible that other factors that may similarly differ between siblings, including factors such as other adversity-related variables (e.g., injuries, school-related stressors) and other relevant challenges (e.g., sleep problems, physical illness). It is possible that the association may in part reflect shared neurobiological or temperament-based vulnerabilities that increase the likelihood of experiencing both bullying victimization and PLEs. For example, genetic variants associated with impairments in neurodevelopment and stress reactivity could contribute to both predisposition to bullying victimization and psychosis spectrum symptoms.(38, 42, 44) Future research should continue to delve into the nature of this bidirectional relationship, including the genetic and neurobiological aspects, between bullying victimization and PLEs.

### Implications

The strong association between distressing PLEs and bullying victimization highlights the importance of early intervention. Preventative strategies may take several forms, including more individual-specific and community-wide strategies. Reducing bullying in schools and community settings is critical, as well as fostering resilience and providing support to bullying victims. These efforts may be broadly beneficial, but may also specifically mitigate the risk of developing PLEs and other related symptoms. Considering that individuals with PLEs are at risk of subsequent bullying victimization, tailored programs that provide psychoeducation and coping mechanisms for youth experiencing PLEs could be beneficial. Schools, mental health professionals, and caregivers who become aware of a youth being victimized should intervene to help amplify stress coping skills to attenuate the negative effects of peer victimization.

### Strengths and Limitations

A strength of the current study is the use of the ABCD Study, a large, demographically diverse, and well-characterized sample that enables the examining of bullying victimization and PLEs over time. In terms of limitations, the present study relied on self-report measures, including youth- and caregiver-reported measures of bullying victimization and youth-reported PLEs. Further, measures of bullying victimization and PLEs do not ask participants when the experience first started, precluding comprehensively modeling whether bullying victimization or PLEs were first to onset. Additionally, the youth-reported measure of bullying victimization is not currently available at every assessment wave (i.e., it is only available at baseline, 1-year, and 2-year follow-up) and is only currently available for a subset of youth, although findings were consistent with caregiver-reported youth bullying victimization experiences.

## Conclusions

Our study provides important evidence for the significant role of bullying victimization in the development and exacerbation of PLEs. Analyses indicate that the association between bullying victimization and PLEs is driven not only by shared factors but also by non-shared factors that are plausibly causal. Specifically, by leveraging longitudinal and within-family designs, our findings suggest that bullying victimization may contribute to PLE risk above and beyond shared familial and genetic influences. These findings have profound implications for preventative and intervention efforts aimed at reducing bullying victimization and supporting vulnerable youth. If this person-specific pathway is causal in nature, it will be critical to institute policies, both public and clinical, to mitigate the negative effects of bullying victimization.

## Supporting information

Supplemental Results

## Data Availability

The ABCD data used in this report came from DOI 10.82525/AB4Q-QR87

## Acknowledgments

Data used in the preparation of this article were obtained from the <u>Adolescent Brain</u> <u>Cognitive Development™ (ABCD) Study</u>, held in the <u>NIH Brain Development Cohorts</u> <u>Data Sharing Platform</u>. This is a multisite, longitudinal study designed to recruit more than 10,000 children aged 9–10 and follow them over 10 years into early adulthood.

The ABCD Study® is supported by the **National Institutes of Health** and additional federal partners under award numbers:

U01DA041048, U01DA050989, U01DA051016, U01DA041022, U01DA051018, U01DA 051037, U01DA050987, U01DA041174, U01DA041106, U01DA041117, U01DA041028, U01DA041134, U01DA050988, U01DA051039, U01DA041156, U01DA041025, U01D A041120, U01DA051038, U01DA041148, U01DA041093, U01DA041089, U24DA0411 23, U24DA041147.

A full list of supporters is available at Federal Partners – ABCD Study.

ABCD Consortium investigators designed and implemented the study and/or provided data but did not necessarily participate in the analysis or writing of this report. This manuscript reflects the views of the authors and may not reflect the opinions or views of the NIH or ABCD Consortium investigators.

The ABCD data repository grows and changes over time. The ABCD data used in this report came from [NIMH Data Archive Digital Object Identifier (DOI)]. DOIs can be found at **<u>DOI 10.82525/AB4Q-QR87</u>**

## References

1. L. Staines, et al., Psychotic experiences in the general population, a review; definition, risk factors, outcomes and interventions. Psychol. Med. 52, 3297 (2022).

2. L. C. Johns, J. van Os, The continuity of psychotic experiences in the general population. Clin Psychol Rev 21, 1125–1141 (2001).

3. N. R. Karcher, Psychotic-like experiences in childhood and early adolescence: Clarifying the construct and future directions. Schizophr. Res. 246, 205–206 (2022).

4. I. Kelleher, et al., Prevalence of psychotic symptoms in childhood and adolescence: a systematic review and meta-analysis of population-based studies. Psychol Med 42, 1857–1863 (2012).

5. N. R. Karcher, et al., Assessment of the Prodromal Questionnaire-Brief Child Version for Measurement of Self-reported Psychoticlike Experiences in Childhood. JAMA Psychiatry 75, 853–861 (2018).

6. N. R. Karcher, et al., Replication of Associations with Psychotic-like Experiences in Middle Childhood from the Adolescent Brain Cognitive Development (ABCD) study. Schizophr. Bull. Open 1, sgaa009 (2020).

7. H. L. Fisher, et al., Specificity of childhood psychotic symptoms for predicting schizophrenia by 38 years of age: a birth cohort study. Psychol Med 43, 2077–2086 (2013).

8. R. Poulton, et al., Children’s Self-Reported Psychotic Symptoms and Adult Schizophreniform Disorder: A 15-Year Longitudinal Study. Arch. Gen. Psychiatry 57, 1053–1058 (2000).

9. J. van Os, R. J. Linscott, I. Myin-Germeys, P. Delespaul, L. Krabbendam, A systematic review and meta-analysis of the psychosis continuum: evidence for a psychosis proneness-persistence-impairment model of psychotic disorder. Psychol Med 39, 179–195 (2009).

10. R. J. Linscott, J. van Os, An updated and conservative systematic review and meta-analysis of epidemiological evidence on psychotic experiences in children and adults: on the pathway from proneness to persistence to dimensional expression across mental disorders. Psychol Med 43, 1133–1149 (2013).

11. N. R. Karcher, et al., Cognitive and Global Morphometry Trajectories as Predictors of Youth Persistent Distressing Psychotic-Like Experiences. Nature. Mental health 3, 1012–1019 (2025).

12. L. R. Steenkamp, et al., Peer-reported bullying, rejection and hallucinatory experiences in childhood. Acta Psychiatr. Scand. 143, 503–512 (2021).

13. G. P. Strauss, I. M. Raugh, V. A. Mittal, B. E. Gibb, M. E. Coles, Bullying victimization and perpetration in a community sample of youth with psychotic like experiences. Schizophr Res 195, 534–536 (2018).

14. A. Braun, et al., Bullying in clinical high risk for psychosis participants from the NAPLS-3 cohort. Soc. Psychiatry Psychiatr. Epidemiol. 57, 1379–1388 (2022).

15. L. H. Chen, T. Toulopoulou, Pathways linking school bullying and psychotic experiences: Multiple mediation analysis in Chinese adolescents and young adults. Front. Psychiatry 13, 1007348 (2022).

16. T. Singham, et al., Concurrent and Longitudinal Contribution of Exposure to Bullying in Childhood to Mental Health: The Role of Vulnerability and Resilience. JAMA Psychiatry 74, 1112–1119 (2017).

17. D. S. Van Dam, et al., Childhood bullying and the association with psychosis in non-clinical and clinical samples: a review and meta-analysis. Psychol. Med. 42, 2463–2476 (2012).

18. D. Stanyon, et al., The role of bullying victimization in the pathway between autistic traits and psychotic experiences in adolescence: Data from the Tokyo Teen Cohort study. Schizophr. Res. 239, 111–115 (2022).

19. M. L. C. Campbell, A. P. Morrison, The relationship between bullying, psychotic-like experiences and appraisals in 14–16-year olds. Behaviour Research and Therapy 45, 1579–1591 (2007).

20. J. P. Selten, E. Van Der Ven, B. P. F. Rutten, E. Cantor-Graae, The Social Defeat Hypothesis of Schizophrenia: An Update. Schizophr. Bull. 39, 1180–1186 (2013).

21. A. Schreier, et al., Prospective study of peer victimization in childhood and psychotic symptoms in a nonclinical population at age 12 years. Arch Gen Psychiatry 66, 527–536 (2009).

22. H. Garavan, H. Bartsch, K. Conway, … A. D.-D. cognitive, undefined 2018, Recruiting the ABCD sample: Design considerations and procedures. Elsevier.

23. N. R. Karcher, D. M. Barch, The ABCD study: understanding the development of risk for mental and physical health outcomes. Neuropsychopharmacology 2020 46:1 46, 131–142 (2020).

24. D. M. Barch, et al., Demographic and mental health assessments in the adolescent brain and cognitive development study: Updates and age-related trajectories. Dev. Cogn. Neurosci. 52, 101031 (2021).

25. N. R. Karcher, J. Schiffman, D. M. Barch, Environmental Risk Factors and Psychotic-like Experiences in Children Aged 9–10. J. Am. Acad. Child Adolesc. Psychiatry 60, 490–500 (2021).

26. N. R. Karcher, et al., Psychotic-like Experiences and Polygenic Liability in the Adolescent Brain Cognitive Development Study. Biol. Psychiatry Cogn. Neurosci. Neuroimaging 7, 45–55 (2022).

27. E. Zoupou, N. R. Karcher, J. J. Jackson, D. M. Barch, Latent multimodal profiles associated with psychosis-like experiences at follow-up. Biol. Psychiatry Cogn. Neurosci. Neuroimaging (2025). 10.1016/J.BPSC.2025.10.017.

28. L. Townsend, et al., Development of Three Web-Based Computerized Versions of the Kiddie Schedule for Affective Disorders and Schizophrenia Child Psychiatric Diagnostic Interview: Preliminary Validity Data. J. Am. Acad. Child Adolesc. Psychiatry 59, 309–325 (2020).

29. J. Kaufman, et al., “KSADS-PL” (Yale University, 2013).

30. R. W. Stewart, C. F. Drescher, D. J. Maack, C. Ebesutani, J. Young, The Development and Psychometric Investigation of the Cyberbullying Scale. J. Interpers. Violence 29, 2218–2238 (2014).

31. A. De Los Reyes, M. J. Prinstein, Applying depression-distortion hypotheses to the assessment of peer victimization in adolescents. J. Clin. Child Adolesc. Psychol. 33, 325–335 (2004).

32. T. M. Achenbach, S. H. McConaughy, M. Y. Ivanova, L. A. Rescorla, Manual for the ASEBA brief problem monitor (BPM). Burlington, VT: ASEBA 1–33 (2011).

33. Y. Rosseel, lavaan: An R Package for Structural Equation Modeling. J. Stat. Softw. 48, 1–36 (2012).

34. D. Pagliaccio, et al., Shared Predisposition in the Association Between Cannabis Use and Subcortical Brain Structure. JAMA Psychiatry 72, 994–1001 (2015).

35. N. R. Karcher, et al., Genetic Predisposition vs Individual-Specific Processes in the Association Between Psychotic-like Experiences and Cannabis Use. JAMA Psychiatry 76, 87–94 (2019).

36. J. M. Boden, S. Van Stockum, L. J. Horwood, D. M. Fergusson, Bullying victimization in adolescence and psychotic symptomatology in adulthood: evidence from a 35-year study. Psychol. Med. 46, 1311–1320 (2016).

37. M. T. McKay, et al., Childhood trauma and adult mental disorder: A systematic review and meta-analysis of longitudinal cohort studies. Acta Psychiatr. Scand. 143, 189–205 (2021).

38. I. Myin-Germeys, J. van Os, Stress-reactivity in psychosis: Evidence for an affective pathway to psychosis. Clin. Psychol. Rev. 27, 409–424 (2007).

39. R. van Winkel, N. C. Stefanis, I. Myin-Germeys, Psychosocial stress and psychosis. A review of the neurobiological mechanisms and the evidence for gene-stress interaction. Schizophr Bull 34, 1095–1105 (2008).

40. A. E. Cullen, S. Rai, M. S. Vaghani, V. Mondelli, P. McGuire, Cortisol responses to naturally occurring psychosocial stressors across the psychosis spectrum: A systematic review and meta-analysis. Front. Psychiatry 11, 1–18 (2020).

41. D. Turley, R. Drake, E. Killackey, A. R. Yung, Perceived stress and psychosis: The effect of perceived stress on psychotic like experiences in a community sample of adolescents. Early Interv. Psychiatry 13, 1465–1469 (2019).

42. R. Mizrahi, Social Stress and Psychosis Risk: Common Neurochemical Substrates? Neuropsychopharmacology 2016 41:3 41, 666–674 (2015).

43. K. Aberizk, et al., Life Event Stress and Reduced Cortical Thickness in Youth at Clinical High-Risk for Psychosis and Healthy Youths. Biol. Psychiatry Cogn. Neurosci. Neuroimaging (2021).

44. D. Mayo, et al., The role of trauma and stressful life events among individuals at clinical high risk for psychosis: A review. Front. Psychiatry 8, 247326 (2017).

