## Supplemental Results for "Disentangling Environmental and Genetic Influences on Associations Between Childhood Bullying Victimization and Psychotic-Like Experiences"

| Supplemental Table S1. PQ-BC Questions and Domain Type | | |
| --- | --- | --- |
| Question Number | Modified PQ-BC Question | PQ-BC Question Type |
| 1 | Did places that you know well, such as your bedroom, or other rooms in your home, your classroom or school yard, suddenly seem weird, strange or confusing to you; like not the real world? | UTC |
| 2 | Did you hear strange sounds that you never noticed before like banging, clicking, hissing, clapping, or ringing in your ears? | PD |
| 3 | Did things you looked at seem different than they usually do; like did they seem shinier or darker, larger or smaller or changed in some other way? | PD |
| 4 | Did you feel like you had special, unusual powers like you could make things happen by magic, or that you could magically know what was inside another person's mind, or magically know what was going to happen in the future when other people could not? | UTC |
| 5 | Did you feel that someone else, who is not you, has taken control over the private, personal, thoughts or ideas inside your head? | UTC |
| 6 | Did you suddenly find it hard to figure out how to say something quickly and easily so that other people would understand what you meant? | DS |
| 7 | Did you ever feel very certain that you have very special abilities or magical talents that other people do not have? | UTC |
| 8 | Did you suddenly feel that you could not trust other people because they seemed to be watching you or talking about you in an unfriendly way? | S |
| 9 | Did your skin or just beneath your skin suddenly start feeling strange, like bugs crawling? | PD |
| 10 | Did you lose concentration because you noticed sounds in the distance that you usually don't hear? | PD |
| 11 | Although you could not see anything or anyone, did you suddenly start to feel that an invisible energy, creature, or some person was around you? | UTC |
| 12 | Did you start to worry at times that your mind was trying to trick you or was not working right? | UTC |
| 13 | Did you feel that the world is not real, you are not real, or that you are dead? | UTC |
| 14 | Did you feel confused because something you experienced didn't seem real or it seemed imaginary to you? | UTC |
| 15 | Did you honestly believe in things that other people would say are unusual or weird? | UTC |
| 16 | Did you feel that parts of your body had suddenly changed or worked differently than before; like your legs had suddenly turned to something else or your nose could suddenly smell things you'd never actually smelled before? | PD |
| 17 | Did you feel that sometimes your thoughts were so strong you could almost hear them, as if another person, NOT you, spoke them? | UTC |
| 18 | Did you feel that other people might want something bad to happen to you or that you could not trust other people? | S |
| 19 | Did you suddenly start to see unusual things that you never saw before like flashes, flames, blinding light, or shapes floating in front of you? | PD |
| 20 | Did you suddenly start to be able to see things that other people could not see or they did not seem to see? | PD |
| 21 | Did you suddenly start to notice that people sometimes had a hard time understanding what you were saying, even though they used to understand you well? | DS |
| *Note.* UTC=unusual thought content ; PD=perceptual distortions; S = suspiciousness; DS=disorganized speech | | |

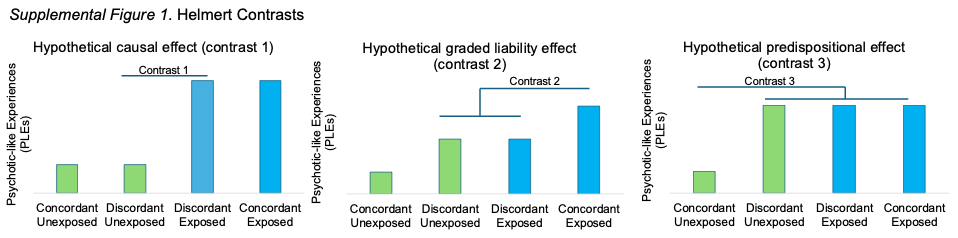

Hypothetical patterns of results from the linear mixed model analyses to test the causal, grade, and predispositional contrasts.

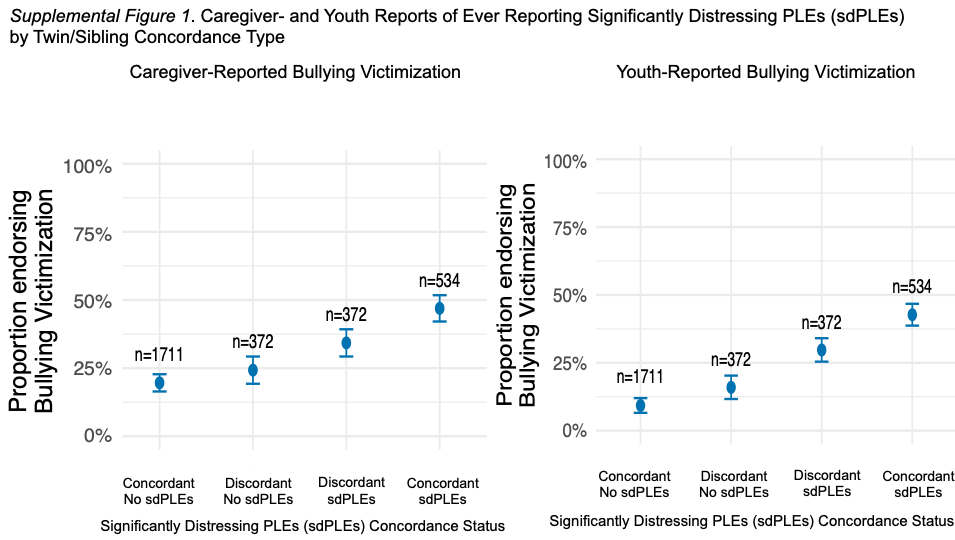

Proportion (and standard error) ever experiencing bullying victimization, as reported by either caregiver or youth, among each of the significantly distressing PLEs concordance types (ie, concordant exposed, discordant exposed, discordant unexposed, concordant unexposed) for twin/sibling pairs.
